# Impact of the COVID-19 pandemic on the mycoplasma pneumoniae infections in children:a retrospective study at a tertiary hospital of Middle China

**DOI:** 10.1101/2025.04.25.25326390

**Authors:** Pengbo Guo, Zhi Lei, Xuan Zheng, Yaodong Zhang, Zhiyi Xia

## Abstract

**Aim:** Mycoplasma pneumoniae (MP) is a common and significant pathogen prevalent among children. This study aimed to investigate the epidemiological characteristics of MP infection in children prior to (2018-2019), during (2020-2022), and following (2023-2024) the COVID-19 pandemic.

**Methods:** This study collected 140,098 MP detection results from Henan Children’ s Hospital from 2018 to 2024. An analysis of the epidemiological characteristics of MP infection in children was conducted.

**Results:** Our findings revealed that the highest positive rate of MP infection reached 35.86% in 2023. Among children diagnosed with pneumonia, the positivity rate was the highest. Additionally, children aged over 6 years had the highest positive rate of MP infection. Furthermore, girls were found to be more susceptible to MP infection than boys. Before the COVID-19 pandemic, a high positivity rate epidemic began in July each year and lasted for over four months. During the pandemic, there was no prolonged seasonal high positivity rate. Post-pandemic, a sharp surge in positive rates began in July, sustaining an intense outbreak for 17 consecutive months, underscoring significant shifts in MP’s epidemiological patterns.

**Conclusions:** The implementation of strict pandemic control measures led to a disruption in the circulation of *MP*. However, with the termination of these control measures, *MP* has witnessed a more severe explosive growth among children, and has shown an epidemic situation with a longer duration.

## 1 Introduction

Respiratory tract infections are common diseases for human, especially for children. The respiratory tract pathogens mainly include bacteria, viruses, chlamydia and mycoplasma. The most common symptoms they caused was pneumonia. *MP* account for 10%-40% of the community-acquired pneumonia (CAP) in children[1].*MP*, recognized as the smallest independent living microorganism, possesses a unique biological status. It is highly contagious, primarily spreading through respiratory droplets generated by coughing and sneezing. The clinical manifestations of MP infection are diverse, commonly presenting as fever, wheezing, cough with expectoration, and in some cases, nosebleeds[2]. These symptoms cannot be distinguished clearly from other respiratory virus infections, and their clinical diagnosis is difficult for majority doctors[3]. Global *MP* epidemics occur every 3–7 years with various incidence rates[4], with some progressing to gastrointestinal, hematological, neurological, cardiovascular, dermatological, and liver system dysfunction, resulting in death in some cases. Therefore, monitoring the epidemiological characteristics of *MP* is important.

In this study, based on the clinical data of Henan Children’s Hospital from January 2018 to December 2024, we conducted a comprehensive retrospective analysis of the positive rate of *MP*. By systematically exploring the epidemic characteristics and distribution patterns of *MP* among children with acute respiratory tract infections, we deeply analyzed its epidemiological features. The research findings are of great significance for optimizing the prevention and control strategies of *MP* infections in children. It is expected that through early intervention measures, the incidence risk of *Mycoplasma pneumoniae* pneumonia (MPP) in children can be significantly reduced, providing more effective protection for children’s respiratory health.Given the potential severity of *MP* infection and its impact on public health, monitoring its epidemiological characteristics is of paramount importance. It aids in the development of effective prevention and control strategies and enhances our preparedness against future outbreaks.

## 2 Materials and methods

### 2.1 Data collection

Children with acute respiratory tract infection in outpatient and inpatient departments of Henan Children’s Hospital from January 2018 to December 2024 were enrolled. Data such as age, gender and diagnosis of the patients were obtained from electronic medical system. All enrolled children met the following criteria: (1) body temperature >37.5 °C and had one or more respiratory symptoms (cough, sore throat, wheezing, etc.) and (2) patients aged under 18 years old. The exclusion criteria for this study were as follows: (1) children with incomplete clinical data; (2) with airway abnormalities and abnormal immune systems (3)children with congenital malignant tumor. All of the children were divided into four age groups: 0–12 months, 1–3 years, 3–6 years, and over 6 years old.

### 2.2 MP detection

Throat swab, sputum or bronchoalveolar lavage fluid(BULF) samples were collected from all enrolled patients on admission and stored in RNA preservation solution. *MP* was detected by SAT-*MP* kit (Rendu Biotechnology Co., Ltd, Shanghai, China). Primers were targeted to 16S rRNA: *MP*-1 (5’AATTTAATACGACTCACTATAGGGAGACACCGCTCCACATGAAATTCCAAAACTCCC 3’) and *MP*-2 (5’CGGTAATACATAGGTCGCAAGC3’). The probe was FAM-5’CGGACUAUUAAUCUAGAGUGUGUCCG3’-DABCYL. The assay has a minimum limit of 1000 copies/ml.

### 2.3 Statistical analysis

Raw data were analyzed using SPSS software (version 21.0; IBM Corp., USA). Positive rates among di□erent age groups and mouths were analyzed using the chi-squared test or Fisher’s exact test. Statistical significance was considered when P-value (two-tailed)< 0.05.All the Figures were created using the Matplotlib package in the Python programming language.

## 3 Results

### 3.1 Overall detection information of *MP* infection in children

Samples from a total of 140, 098 children were analyzed from January 2018 to December 2024 in Henan Children’s Hospital, including 15,493 cases in 2018, 23,194 cases in 2019, 13, 629 cases in 2020, 14, 561 cases in 2021, 9, 195 cases in 2022,28, 301 cases in 2023, 35,726 cases in 2024 (Table 1). In Henan, 27, 923 positive cases, 15, 420 (55.23 %) were boys, and 12, 503 (44.77 %) were girls, resulting in a gender ratio of 1:1.23. We analyzed the data of male and female. The overall gender difference from 2018 to 2024 showed that the positive rate in males was significantly lower than females (p < 0.001). Specifically, in 2019, boys had a significantly lower positive rate than girls (p < 0.001), and this trend persisted in 2023 and 2024 (P< 0.01) (Fig 5).Our analysis revealed that throat swabs were the most commonly obtained samples followed by BALF and sputum. (Table 1and Fig 1)

**Table 1.** Basic information of MP detection from 2018 to 2024.

| Characteristic |  | 2018<br>(15,493) | 2019<br>(23,194) | 2020<br>( 13,629) | 2021<br>( 14,561) | 2022<br>(9,195) | 2023<br>(28,301) | 2024<br>(35,726) |
| --- | --- | --- | --- | --- | --- | --- | --- | --- |
| MP positive specimens |  | 1705 | 3249 | 1192 | 1636 | 1117 | 10151 | 8873 |
| Gender | Male | 971 | 1780 | 682 | 956 | 611 | 5569 | 4851 |
|  | Female | 734 | 1469 | 510 | 680 | 506 | 4582 | 4022 |
| Sample types | Throat swab | 545 | 1894 | 789 | 1345 | 974 | 9543 | 8521 |
|  | BALF | 543 | 1132 | 381 | 269 | 132 | 300 | 338 |
|  | Sputum | 617 | 223 | 22 | 22 | 11 | 308 | 14 |

**Fig 1.**
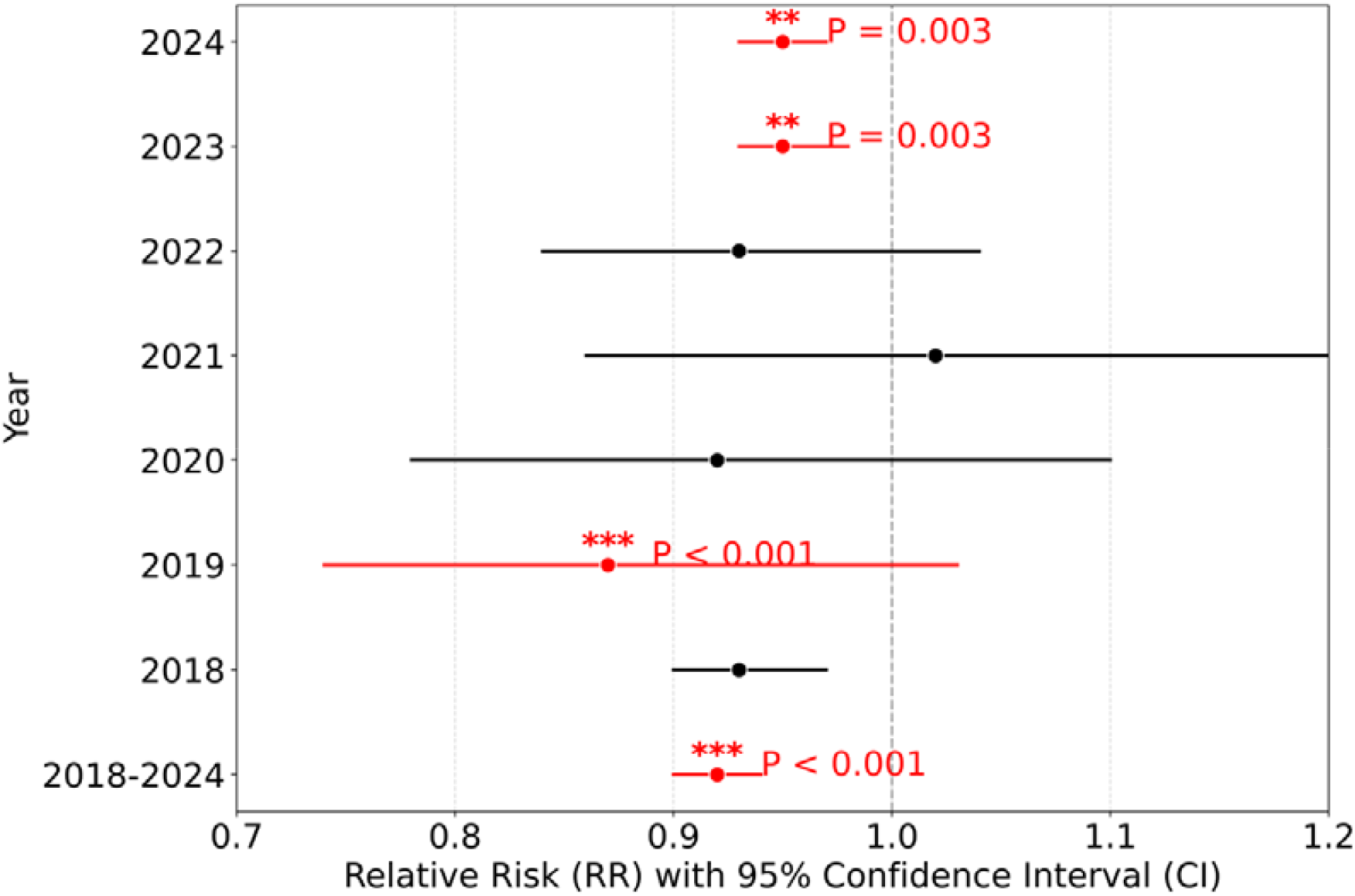
Yearly distribution of MP infection in children.

**Fig 2.**
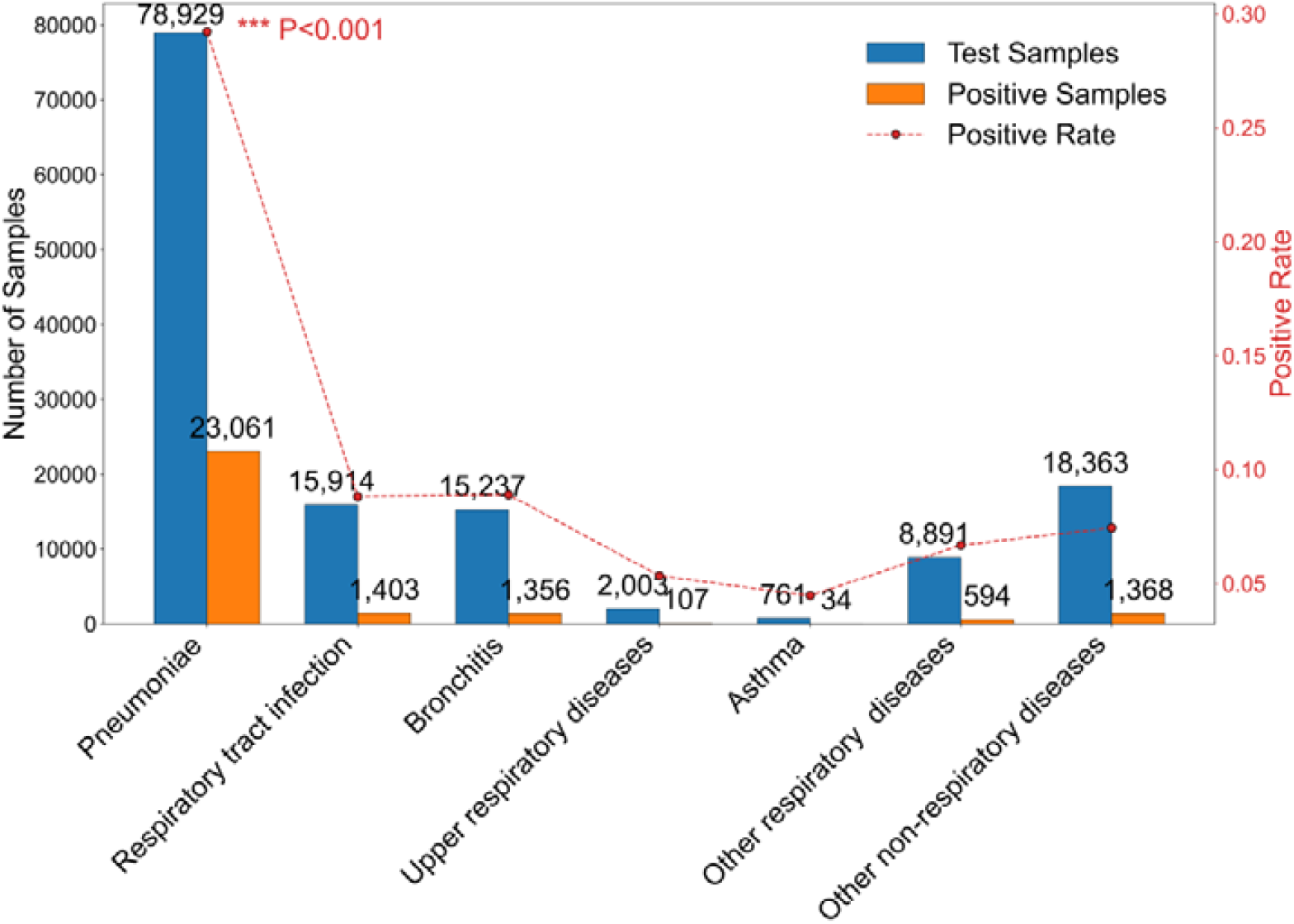
The RR value and P value analysis of male and female.

### 3.2 Clinical manifestations of *MP* infection

*MP* infection with a clear medical diagnosis were analyzed. Respiratory system-associated diagnosis accounted for 86.89 % (12,1735/14, 0098) of cases with a significant difference of positive rate in comparison to other systems (χ2 = 2060.084, p <0.001), and most of these cases were diagnosed as pneumonia caused by *MP*. The types of pneumonia included asthmatic pneumonia, lobar pneumonia, interstitial pneumonia,inhalation pneumonia, wheezing pneumonia, obstructive pneumonia, prolonged pneumonia and bronchopneumonia. Non-pneumonia respiratory manifestations include respiratory tract infections, bronchitis, upper respiratory associated disease (such as rhinitis and pharyngitis), influenza, asthma and other disorders. (Table 2 and Fig 3)

**Table 2.** Clinical manifestations of children with and without MP infection.

| Diagnosis | Total<br>(n=140098) | Positive<br>(n=27923) | positive rate |
| --- | --- | --- | --- |
| Respiratory and infectious diseases | 121735 | 26555 | 21.80% |
| Pneumoniae | 78929 | 23061 | 29.22% |
| Respiratory tract infection | 15914 | 1403 | 8.82% |
| Bronchitis | 15237 | 1356 | 8.90% |
| Upper respiratory tract associated diseases | 2003 | 107 | 5.34% |
| Asthma | 761 | 34 | 4.47% |
| Other respiratory diseases | 8891 | 594 | 6.68% |
| Other non-respiratory diseases | 18363 | 1368 | 7.45% |

\*\*\* P<0.001

**Fig 3.**
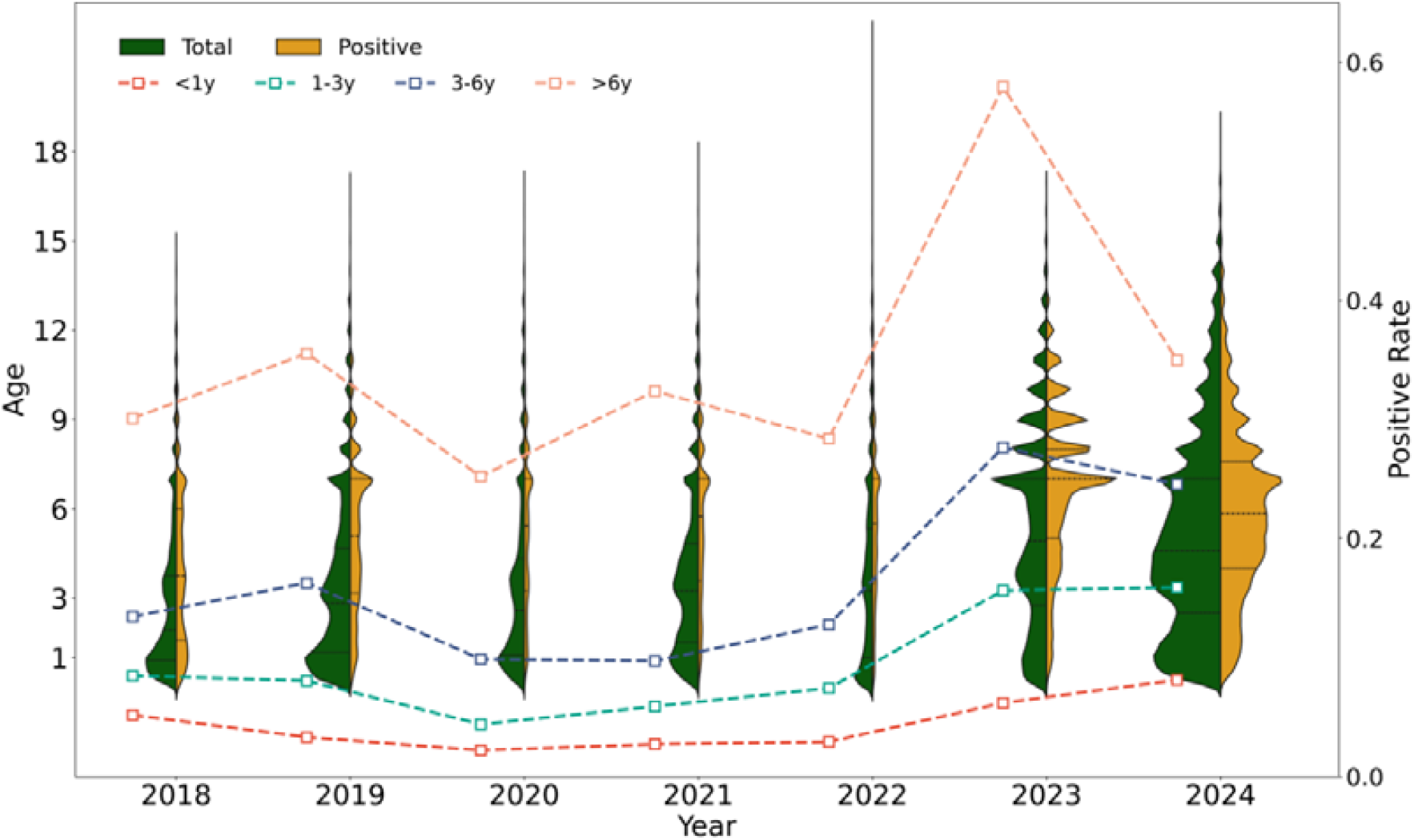
Clinical manifestations of *MP* infection

### 3.3 Age distribution of *MP* infection in children

The mean age of children testing positive for *MP* was 5.91 ± 2.77 years, with a range spanning from 0.06 to 17.00 years. During the 2018-2024 period, two cases of MP infection were identified in infants under one month of age. The age distribution of MP infection is depicted in Fig 4. Both the positivity rate and infection count demonstrated a gradual increase with age, peaking in the over-6-year-old group, where the positivity rate was significantly higher than in all other age groups (P<0.001). Notably, the number of MP-positive children in this older age group was sixfold higher than that in the 0-12 month cohort (3120 vs. 522 cases, respectively). In 2023, the highest positivity rates were recorded in the over-6-year-old and 3–6-year-old groups, highlighting distinct age-related patterns in *MP* infection prevalence.

**Fig 4.**
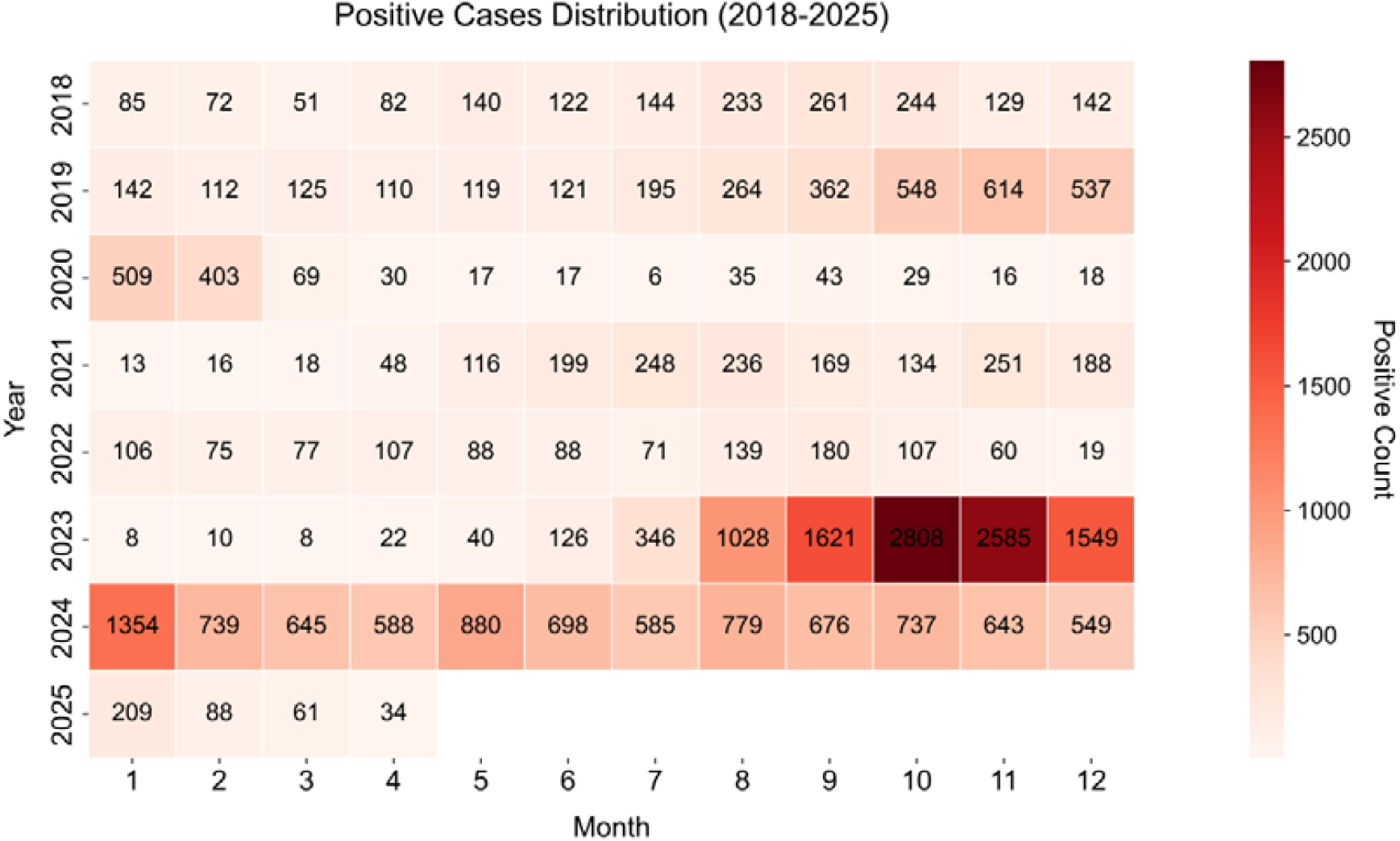
Age distribution of MP infection in children.

**Fig 5.**
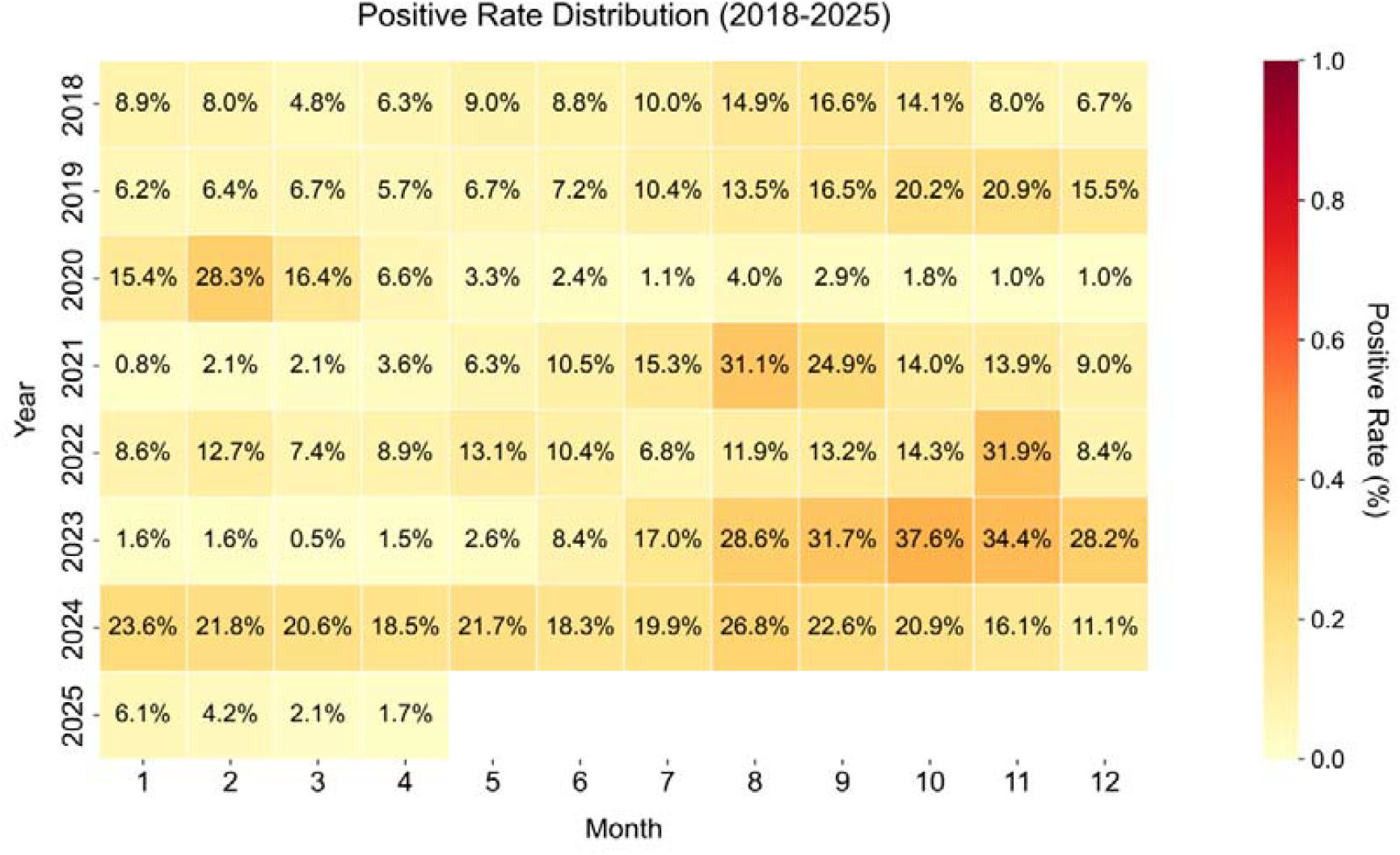
Month distribution of MP infection in children.

### 3.4 Time distribution of *MP* infection in children

Fig 1 illustrates the distribution of the annual number of MP-positive cases and the corresponding positivity rates from 2018 to 2024. On an annual basis, the positivity rate of MP was relatively low before the pandemic, at 11% in 2018 and 14% in 2019. During the three years of the pandemic, the positivity rate remained at a lower level, increasing slightly from 9% in 2020 to 12% in 2022. After the pandemic, the positivity rate surged dramatically to 36% in 2023, which was significantly higher than in any other year over the past seven years (P < 0.001). In 2024, the positivity rate remained high at 25%, compared with the pre-pandemic levels.

A more detailed monthly analysis (Fig 5) reveals that before the pandemic, the MP infection rate was relatively low from January to June (with an average of 7.06%), but it increased significantly from July and remained high for an extended period (with an average positivity rate of 16.36%). During the pandemic, except for August 2021, September 2021, and November 2022, the MP infection rate was generally low, with no sustained high positivity rate for an extended period. After the pandemic, from January to June 2023, the MP positivity rate was low, consistent with the low positivity rate observed during the pre-pandemic period from January to June. However, starting in July 2023, there was a dramatic increase in both the number of MP infections and the positivity rate (with an average positivity rate of 29.68% from July to December). In terms of positive numbers, the total number of MP-positive cases from July to December 2023 (9,591) exceeded the total number of positive cases from 2019 to 2022 (8,899). This high positivity rate persisted for 17 months until November 2024, with an average positivity rate of 20.98% from January to November 2024. The positivity rate began to decline significantly in December 2024. To further summarize the prevailing patterns of MP, data for January to April 2025 (updated to April 20, 2025) were added. The MP positivity rate decreased significantly and gradually during this period, aligning with the low epidemic trend observed from January to June before the pandemic. However, after the prolonged high positivity outbreak, the positivity rate in January to April 2025 was extremely low (with an average of 3.52%).

## 4 Discussion

This study systematically analyzed the epidemiological characteristics of Mycoplasma pneumoniae (MP) infection in children before the COVID-19 pandemic (2018-2019), during the pandemic (2020-2022), and after the pandemic (2023-2024). The results showed that there were significant differences in the distribution of *MP* infection in terms of gender, age, and time, providing an important basis for optimizing clinical diagnosis and treatment as well as public health prevention and control strategies.

The positive rate of *MP* in children with pneumonia was the highest (29.22%, Table 2), once again verifying that *MP* is a core pathogen of community-acquired pneumonia in children. In contrast, the positive rate in children with asthma was the lowest (4.47%), suggesting a weak association between *MP* infection and acute asthma attacks. In clinical practice, for children with pneumonia (especially during the epidemic period after the pandemic), *MP* detection should be given priority to guide the precise treatment with macrolide drugs and reduce the risk of severe illness.

Pre-pandemic, *MP* infection exhibited typical seasonal characteristics, with the positive rate rising sharply from July and lasting for more than four months (Fig 5), which was consistent with the previously reported periodic epidemic pattern of *MP* (3-7 years) and its respiratory droplet transmission characteristics. However, during the pandemic, strict non-pharmaceutical interventions (NPIs), such as wearing masks, social restrictions, and school closures, significantly interrupted the seasonal transmission of *MP*. There was no persistent high positive rate from 2020 to 2022 (Fig.1), indicating that NPIs not only inhibited the transmission of the novel coronavirus but also indirectly weakened the epidemic intensity of *MP*, similar to the observations of other respiratory pathogens such as influenza.

It was reported that during the *MP* epidemics, 20–40 % of pneumonia cases accounted to MP in the general population[5–7].Post-pandemic, MP infection rebounded explosively, with the peak positive rate reaching 35.86% in 2023 (Fig 1), far exceeding the pre-pandemic level. But in the study of Yue, there was no *MP* epidemic in North China in 2023[11].This may be because of the difference of temperature. Japanese studies have suggested that increases had positive correlation with the infection rate of *MP*[9, 10]. This study also found that the epidemics could occur in any season according to the *MP* epidemiological characteristics in 2024.Similar phenomena of “immune debt” have been reported in many places around the world, that is, long-term low exposure to pathogens leads to a decrease in herd immunity, and there is an increase in infections after the pandemic To explain this phenomenon, Li proposed a hypothesis: immune gap or immune debt[8].But it need more study to verify this hypothesis.It is worth noting that, according to the latest data from the first few months of 2025, after a high level of “repayment” of the immune debt, the child population seems to have established a temporarily stable immune barrier, and the positive rate of *MP* has reached the lowest level in recent years (Fig 5). This phenomenon provides new clues for our further research on the immune mechanism of the child population and the epidemic pattern of *MP*, but the long-term change trend still needs to be continuously monitored.

Children may be infected by *MP* at any age. *MP* was the most common pathogenin children aged 6–17 years. This is basically consistent with previous studies[12–14]. Children older than 6 years are children of schoolage who spend more time in the classroom or be centrally managed compare with children under 6 years old. This also proofed the previous reports that MP spread more easily in close contact and hermetic environment[15]. The number of infected children in this group was six times that of the infant group (0-12 months old). In contrast, the positive rate in the infant group was the lowest, which was probably because of their relative independent living environment. Islam’s study shown that breast feeding could reduce the risk of acute respiratory infection in infants[16]. This result suggests that educational institutions such as schools need to strengthen targeted prevention and control measures, such as promoting hand hygiene, respiratory etiquette, and environmental ventilation. In addition, the infection risk of female children was significantly higher than that of male children (p<0.001, Fig 5), which may be related to differences in immune responses, hormone levels, or medical-seeking behaviors. While in the study of Yue, the sex ratio (male to female) of children with *MP* infections was approximately 1.34[11]. Further research on the gender-related susceptibility mechanisms is needed.

### Research limitations and prospects

This study has the following limitations: (1) The data are from a single center in Central China, so caution should be exercised when extrapolating the conclusions; (2) The retrospective design makes it difficult to rule out confounding factors; (3) The epidemic trend of *MP* after 2025 still requires long-term observation. In the future, multi-center studies should be carried out, combined with pathogen genomics and drug resistance monitoring, to deeply analyze the rebound mechanism of *MP* (such as antigenic drift and antibiotic resistance), and explore the feasibility of vaccine development.

## Conclusion

The COVID-19 pandemic temporarily changed the epidemic pattern of *MP* through NPIs, but its sharp rebound after the pandemic revealed the dynamic balance between the adaptability of pathogens and herd immunity. It is recommended to strengthen active monitoring in high-risk seasons, promote vaccine development, and enhance prevention and control awareness through public education. Clinically, attention should be paid to MP-related pneumonia in school-age children. Early diagnosis and intervention are the keys to improving the prognosis.

## Authors’ contributions

PengboGuo and Zhiyi Xia conceived the idea and designed the study. PengboGuo, YaodongZhang, Xuan Zheng and ZhiLeicontributed to the data processing. Xuan Zheng and ZhiLei prepared all tables and figures. PengboGuo and Yaodong Zhang contributed to the statisticalanalysis. All authors wrote and revised the manuscript and approved the manuscript submission.

## Data availability

The datasets used and/or analyzed inthis study are available from the corresponding author (Zhiyi Xia) on request.

## Declarations

## Ethics approval and consent to participate

The study was approved by the Ethics Committees of Henan Children’s Hospital(2023-k-078)

The informed consents of patients were waived by both Ethics Committees due to the retrospective nature of this study.

## References

1. Xue Y, Wang M, Han H (2023) Interaction between alveolar macrophages and epithelial cells during Mycoplasma pneumoniae infection. Front Cell Infect Microbiol 13:1052020. 10.3389/fcimb.2023.1052020

2. Chen Z, Ji W, Wang Y, et al (2013) Epidemiology and associations with climatic conditions of Mycoplasma pneumoniae and Chlamydophila pneumoniae infections among Chinese children hospitalized with acute respiratory infections. Ital J Pediatr 39:34. 10.1186/1824-7288-39-34

3. Yun KW (2024) Community-acquired pneumonia in children: updated perspectives on its etiology, diagnosis, and treatment. Clin Exp Pediatr 67:80–89. 10.3345/cep.2022.01452

4. Guo P, Mei S, Wang Y, et al (2023) Molecular typing of Mycoplasma pneumoniae and its correlation with macrolide resistance in children in Henan of China. Indian Journal of Medical Microbiology 46:100435. 10.1016/j.ijmmb.2023.100435

5. Atkinson TP, Waites KB (2014) Mycoplasma pneumoniae Infections in Childhood. Pediatric Infectious Disease Journal 33:92–94. 10.1097/INF.0000000000000171

6. Loens K, Goossens H, Ieven M (2010) Acute respiratory infection due to Mycoplasma pneumoniae: current status of diagnostic methods. Eur J Clin Microbiol Infect Dis 29:1055–1069. 10.1007/s10096-010-0975-2

7. Jacobs E, Ehrhardt I, Dumke R (2015) New insights in the outbreak pattern of Mycoplasma pneumoniae. International Journal of Medical Microbiology 305:705–708. 10.1016/j.ijmm.2015.08.021

8. Li H, Li S, Yang H, et al (2024) Resurgence of Mycoplasma pneumonia by macrolide-resistant epidemic clones in China. The Lancet Microbe 5:e515. 10.1016/S2666-5247(23)00405-6

9. Onozuka D, Chaves LF (2014) Climate Variability and Nonstationary Dynamics of Mycoplasma pneumoniae Pneumonia in Japan. PLoS ONE 9:e95447. 10.1371/journal.pone.0095447

10. Onozuka D, Hashizume M, Hagihara A (2009) Impact of weather factors on Mycoplasma pneumoniae pneumonia. Thorax 64:507–511. 10.1136/thx.2008.111237

11. Yue Y, Wu D, Zeng Q, et al (2025) Changes in children respiratory infections pre and post COVID-19 pandemic. Front Cell Infect Microbiol 15:1549497. 10.3389/fcimb.2025.1549497

12. Guo P, Wang Y, Zhou M, et al (2021) Detection and Analysis of Respiratory Pathogens in Hospitalized Children with Acute Respiratory Tract Infections by a Multiplex-PCR Assay Based on the Genetic Analyzer Platform. Iran J Pediatr 31:. 10.5812/ijp.110203

13. Jain S, Williams DJ, Arnold SR, et al (2015) Community-Acquired Pneumonia Requiring Hospitalization among U.S. Children. N Engl J Med 372:835–845. 10.1056/NEJMoa1405870

14. Lee K-L, Lee C-M, Yang T-L, et al (2021) Severe Mycoplasma pneumoniae pneumonia requiring intensive care in children, 2010–2019. Journal of the Formosan Medical Association 120:281–291. 10.1016/j.jfma.2020.08.018

15. Steinberg P, White RJ, Fuld SL, et al (1969) ECOLOGY OF MYCOPLASMA PNEUMONIAE INFECTIONS IN MARINE RECRUITS AT PARRIS ISLAND, SOUTH CAROLINA1. American Journal of Epidemiology 89:62–73. 10.1093/oxfordjournals.aje.a120916

16. Islam M, Islam K, Dalal K, Hossain Hawlader MD (2024) In-house environmental factors and childhood acute respiratory infections in under-five children: a hospital-based matched case-control study in Bangladesh. BMC Pediatr 24:38. 10.1186/s12887-024-04525-4

